# Knowledge, attitudes, and perceptions of the Greek population regarding the COVID-19 pandemic during the national lockdown (March 23 - May 03, 2020): A web-based cross-sectional study

**DOI:** 10.1101/2020.12.19.20248550

**Authors:** Andreas Anestis, Odysseas Lomvardeas, Nikolaos Papadakis

## Abstract

**Objective:** The study assessed the knowledge, attitudes, and perceptions toward the COVID-19 pandemic during the total lockdown of spring 2020 in Greece.

**Methods:** A web-based cross-sectional study was conducted from April 13 to May 5, 2020. Adult residents of Greece anonymously completed an online survey that was distributed through email and social media. A total of 1396 fully complete questionnaires were collected. Demographic questions, questions regarding the knowledge about the disease etiology, diagnosis and prevention, and questions related to the respondents’ attitude and perception toward the restriction measures and the confidence in different kinds of institutions providing information were included in the survey items. The appropriate statistical analyses were conducted according to the type of variable and the research question.

**Results:** The highest knowledge scores were found in females (74.8%, p = 0.015), individuals over 60 years old (77.3%, p < 0.001) and individuals having completed post-secondary or tertiary education (76.5%, p < 0.001). Five attitude patterns toward the pandemic were identified: “trust in institutions”, “trust in the restriction measures”, “trust in media and the internet”, “trust in traditional institutions”, and “measures deniers”. Age, education, and knowledge score were the factors defining the attitudinal patterns revealed.

**Conclusions:** Education and public awareness of scientifically accurate information are the means for eliminating individually and socially irresponsible and dangerous behaviors and protecting public health in periods of health crises.

## 1. INTRODUCTION

Coronavirus disease 2019 (COVID-19), having emerged in Wuhan City, Hubei Province, China, has been declared as a global public health emergency by the World Health Organization (WHO) on January 30, 2020 [1]. After the report of an enormous number of new cases across 118 countries during the first week of March 2020, the disease was characterized as a pandemic [2]. Human - to - human viral transmission through droplet and direct contact have been identified as the main ways through which COVID-19 is rapidly spread. The lack of antiviral treatment or vaccine during the first months of the novel health crisis highlighted the significance of applying the appropriate prevention measures to control infections [3].

The first confirmed COVID-19 case in Greece was reported on February 26, 2020. On the same day, the public information campaign about the disease by the Greek General Secretariat for Civil Protection, titled “We do not fear, Knowledge is our shield,” was launched. The subsequent increase in the number of confirmed cases and deaths led to a progressive suspension of the operation of educational institutions (March 10), theaters and restaurants (March 13) and religious ceremonies (March 16). Finally, the Greek Government implemented a total lockdown on March 22, 2020. During the lockdown period, a daily press conference by the scientific representative of the Greek Ministry of Health, and the under-secretary, responsible for Civil Protection and Crisis Management was taking place, informing the public about the disease progression both in Greece and globally. During the same period, a new campaign titled “We stay at home, we stay safe” was launched [4]. The lifting of the lockdown restrictions occurred gradually, being initiated on May 4, 2020. Greece’s management of the first wave of the pandemic has been characterized as successful and the country has been described as an example of handling the first wave of the health crisis, showing the significance of acting quickly and informing people about the severity of the threat [5].

The pandemic has globally caused not only the loss of numerous human lives, but also job losses, and deterioration of healthcare services and national economies [6]. In such periods of crisis, maintaining social integrity and functions depends on the level of public knowledge, the type of attitude and behavior toward the applied policies and measures and the confidence in authorities and institutions. Surveying knowledge, attitudes and perceptions allows information on what is known, believed, and done by the members of a population, to be collected. The analysis of such data can reveal the factors underlying possible trends and defining the major patterns of individual and social responses against the pandemic.

To the best of our knowledge, the present study is the first assessing the knowledge, attitudes, and perceptions of the adult Greek population toward the COVID-19 pandemic during the total lockdown period (March 22 – May 03, 2020), to shed light on the parameters having shaped the success of the “Greek example”. Additionally, we investigated whether the observed trends are organized into patterns within the study population and to identify the major factors defining these patterns.

## 2. MATERIALS AND METHODS

### 2.1 Study design and sample size

A cross-sectional, web-based questionnaire study was conducted from April 13 to May 5, 2020. The design and reporting of the study adhered to the STROBE guidelines for cross-sectional studies in epidemiology [7]. Snowball sampling was selected because of the difficulty in accessing subjects during the total lockdown, due to the imposed precautionary measures against COVID-19. Greek adults living in Greece during the period the study was conducted, were the target population. The sample size was calculated using the Raosoft sample size calculator [8]. Given the size of the Greek population and for a response distribution of 50% the minimum recommended sample size was 664 (99% confidence level, 5% margin of error).

### 2.2 Survey Development and Distribution

The survey instrument was in Greek and consisted of 49-items divided into three sections: demographics, COVID-19 related knowledge and self-reported attitude and perception. The instrument was developed using a W.H.O. course material on COVID-19 methods for detection, prevention, response, and control [9]. Before the questionnaire was administered, an internal consistency test was conducted in a pilot study among 45 randomly chosen participants and Cronbach’s alpha values for the Knowledge and Attitude - Perception sections were calculated. The time required to answer the survey was also assessed. The results of the pilot study were excluded from the final analysis. For assessing clarity and readability, the questionnaire was reviewed by 2 faculty members. Their suggestions for refinements were adopted in the final version of the instrument.

The survey was administered using Google Forms. Researchers’ email and social networks contacts, as well as participants of online and open fora and groups were sent the link for the questionnaire. The responses were automatically entered into a Google database. Before data entry, participants were informed about the average length of time of the survey, the type of data stored, the database and the duration of their storage, the identity of the researchers and the purpose of the study.

### 2.3 Content of the Survey Instrument

The 49-item questionnaire consisted of closed-ended questions and it comprised three sections. The first section assessed the following demographic characteristics of the participants: gender, age, educational level, and prefecture of residence. The second section included 23 items and assessed the respondents COVID-19 related knowledge regarding the disease etiology, diagnosis, transmission, symptoms, prevention, and treatment. A scoring system was used for the questions of this section. Correct answers were scored 1 point and incorrect or “I do not know” answers were scored 0 points. The overall knowledge scores were calculated from the total sum of correct responses and they were converted into percentiles. The third section of the questionnaire assessed the attitudes and perceptions of the respondents toward the pandemic. The items of this section were organized into two parts. The first part (12 items) was asking for the degree of agreement to statements regarding the significance, efficiency and impact of the restriction measures, and practices related to the prevention against COVID-19. The second part (10 items) assessed the confidence of the participants in political, scientific, social and religious institutions as information providers. All the responses of the third part were recorded on a 1 to 5-point Likert scale (“very low”, “very high”). It is clarified that, throughout the text, trust and confidence are used as synonymous terms.

### 2.4 Data Analysis

The data obtained were coded, validated, and analyzed using Microsoft Excel 2019 and Jamovi version 1.6.8 (The Jamovi project (2020)). In the analysis only completed questionnaires were used. Descriptive analysis was applied for calculating the frequencies and proportions for the categorical variables and the central tendency (mean, median) and dispersion (standard deviation, interquartile range – IQR) for the numerical variables, as appropriate.

The Kolmogorov-Smirnov normality test was applied for assessing the normality of the distribution of continuous variables; the role of demographics in the knowledge, attitude and perception of the respondents was investigated by comparing the differences in the responses of demographic groups using parametric or non-parametric analysis of variance (ANOVA), as appropriate. In all comparisons, a p value of less than .05 was considered statistically significant. Moreover, pairwise comparisons were conducted to identify which pairs of levels of the demographic variables significantly differ from each other.

Additionally, multiple linear regression was used to identify the main demographic variables (predictors) that define the COVID-19 related knowledge scores. A full model (all interactions included) was initially constructed, before the predictor with the largest p value (p>0.05) was removed. The final model included only the predictors that were statistically significant (p < 0.05). Collinearity checks for the identification of interactions between the independent variables were also conducted.

Principal component analysis (PCA) was performed for all attitude/perception items of the questionnaire. The technique was used to reduce the number of variables and to reveal patterns of association between them. The Keiser – Meyer – Olkin (KMO) measure of sampling adequacy and Bartlett’s sphericity test were applied prior to the PCA to show whether associations between the original variables exist. The orthogonal varimax rotation was used to increase the interpretability of the produced factors (components). Naming of the factors and the respective patterns of attitude/perception regarding COVID-19 was based on the factor loadings, considering the highest loadings. Only factor loadings equal or higher than 0.2 (or lower than - 0.2) were considered in the interpretation of the pattern. Solutions of 2 – 5 factors were tested to decide on the factors that should be retained. After the PCA, associations between attitude/perception patterns and knowledge scores were assessed through linear regression modeling, using the attitude and perception items per principal component as the predictors.

### 2.5 Ethical Considerations

Participants’ information remained anonymous and confidentiality of personal information was protected throughout the study. The participation was voluntary, and the respondents were asked to provide honest answers. An informed consent was included on the initial page of the survey. The study was conducted in accordance with the ethical standards of the Declaration of Helsinki as revised in 2013 and had the Approval of the Bioethics Committee of the Medical School of Aristotle University.

## 3. RESULTS

### 3.1 Instrument validity and completeness rate

The internal consistency of the survey was confirmed using the results of the pilot study. Cronbach’s alpha values for the knowledge and attitude - perception sections were 0.76 and 0.71, respectively. The overall Cronbach’s alpha value was 0.72. A total of 1537 individuals responded to the questionnaire and 1396 of them were completed without blanks, yielding a completeness rate of 90.82%.

### 3.2 Study population characteristics and knowledge scores

Participant characteristics regarding gender, age and education are shown in Table 1. Of the total number of participants, 64.6% were female, and most were in the 21 - 29 age class (31.1%). Almost half of the participants (48.5%) had a post-secondary diploma or a university degree. All prefectures were represented in the study population and the distribution of the participants in them was similar to the distribution of the general population (data not shown).

**Table 1.** Size and knowledge scores of the demographic groups of the study population. Comparisons within study subgroups with the Kruskal Wallis nonparametric test (N=1396).

| Variable | Level | Size of subgroups | | Knowledge Score<br>(0-100) | | p | $\epsilon^2$ |
| --- | --- | --- | --- | --- | --- | --- | --- |
|  |  | n | % | Median | IQR |  |  |
| Gender | Male | 494 | 35.4 | 73.9 | 68.1, 78.2 | 0.015* | 0.004 |
|  | Female | 902 | 64.6 | 74.8 | 69.7, 79.0 |  |  |
| Age (years) | 18 - 20 | 175 | 12.5 | 71.4 | 67.2, 75.6 | <0.001*** | 0.045 |
|  | 21 - 29 | 435 | 31.1 | 73.9 | 68.2, 77.3 |  |  |
|  | 30 - 39 | 235 | 16.8 | 74.8 | 69.7, 79.0 |  |  |
|  | 40 - 49 | 266 | 19.1 | 75.6 | 71.4, 79.8 |  |  |
|  | 50 - 59 | 191 | 13.7 | 75.6 | 71.4, 79.0 |  |  |
|  | 60 and over | 94 | 6.7 | 77.3 | 70.8, 79.0 |  |  |
| Education Level<br>(highest completed) | Primary, and secondary | 719 | 51.5 | 73.8 | 68.1, 77.3 | <0.001*** | 0.078 |
|  | Post - secondary and tertiary | 677 | 48.5 | 76.5 | 73.1, 79.0 |  |  |

\*significant at $p < 0.05$ , \*\*significant at $p < 0.01$ , \*\*\*significant at $p < 0.001$ , $\epsilon^2$ : effect size

The results of the Kolmogorov - Smirnov normality test indicated that the knowledge scores in the study population were not normally distributed (p < 0.01). The median of the knowledge scores was 74.8%. Statistically significant differences in the knowledge scores were identified between genders, ages and education levels (Table 1). The highest median knowledge scores were found in females (74.8%, p = 0.015), individuals over 60 years old (77.3%, p < 0.001) and individuals having completed post-secondary or tertiary education (76.5%, p < 0.001). The overall effect size indicated weak - moderate associations between the variables, the strongest being the one between the educational level and knowledge score (ε^2^ = 0.078).

Linear regression modelling was used to further investigate the combined effect of the demographic variables on the knowledge score. Collinearity checks (VIF and tolerance) confirmed the lack of interaction between the independent variables (age, gender, education level and prefecture). Linear regression confirmed the weak association between the independent variables and the knowledge score (R^2^ = 0.125). Moreover, the prefecture was the only non-significant factor (p > 0.05) (Table 2), thus it was excluded from the final model (Table 3).

**Table 2.** Results of the omnibus ANOVA test for the overall fit of the linear model. Statistically significant p values are in bold.

| Demographic variable | F | p |
| --- | --- | --- |
| Gender | 19.972 | **<0.001** |
| Age | 6.307 | **<0.001** |
| Educational level | 6.263 | **<0.001** |
| Prefecture | 0.903 | 0.544 |

**Table 3.** Final demographic model for the knowledge score.

| Coefficient | Estimate | SE | p value |
| --- | --- | --- | --- |
| Intercept | 72.27 | 6.93 | <0.001*** |
| Female | 1.76 | 0.39 | <0.001*** |
| 21-29 y.o. | 0.82 | 0.74 | 0.271 |
| 30-39 y.o. | 2.43 | 0.85 | 0.005** |
| 40-49 y.o. | 3.74 | 0.83 | <0.001*** |
| 50-59 y.o. | 3.86 | 0.86 | <0.001*** |
| 60 y.o. and over | 3.88 | 1.02 | <0.001*** |
| Tertiary education | 3.33 | 0.58 | <0.01** |

*Reference group: male, 18-20 y.o., having completed the primary or secondary education.*
\*significant at $p < 0.05$ , \*\*significant at $p < 0.01$ , \*\*\*significant at $p < 0.001$

### 3.3 Attitudes and perceptions

Attitudes and perceptions of the study population toward COVID-19 pandemic were recorded as the degree of agreement to a set of statements (Table 2) and the level of confidence in different institutions (Table 3). The highest mean agreement scores were recorded for adopting the precautionary measures against COVID-19 (4.18 ± 0.85) and the perception that the viral spread will be eliminated due to the restriction measures (3.82 ± 1.59).

Medical and scientific institutions seemed to have gained the trust of the public (level of confidence range: 3.66 - 3.92), while this was not true for political and religious institutions and social networks (level of confidence range: 1.52 - 1.90).

Statistically significant differences in the attitudes and perceptions of the respondents were identified mainly between ages and education levels (Tables 4, 5). The results of the pairwise comparisons between levels of the demographic variables (data not shown) are argued in the Discussion session.

**Table 4.** Respondents agreement with statements regarding attitudes and perceptions toward the COVID-19 pandemic. Comparisons within study subgroups with the Kruskal Wallis nonparametric test (N=1396).

| Overall degree of agreement and significance of sub-groups differences |  |  |  |  |  |  |  |
| --- | --- | --- | --- | --- | --- | --- | --- |
| Statements | Overall Agreement (M ± SD) | Gender differences |  | Age differences |  | Educational level differences |  |
|  |  | p | ε<sup>2</sup> | p | ε<sup>2</sup> | p | ε<sup>2</sup> |
| Confirmed COVID-19 cases that violate the restriction measures should pay at least partially for their treatment | 2.77 ± 1.55 | 0.003** | 0.006 | 0.062 | 0.009 | 0.954 | 0.006 |
| Violating the restriction measures is unethical | 3.42 ± 1.60 | 0.223 | 0.001 | 0.055 | 0.009 | 0.187 | 0.016 |
| The economic impacts of the lockdown will be tremendous | 3.46 ± 1.43 | 0.329 | 0.000 | 0.023* | 0.010 | 0.037* | 0.020 |
| The Public Healthcare System can cope with the COVID-19 outbreak | 2.74 ± 1.26 | 0.293 | 0.000 | 0.001** | 0.016 | 0.420 | 0.012 |
| Restriction measures had to be adopted regardless the economic impact | 3.45 ± 1.62 | 0.131 | 0.002 | 0.001** | 0.016 | 0.141 | 0.017 |
| Restriction measures violate human rights | 2.28 ± 1.34 | 0.414 | 0.000 | 0.439 | 0.004 | 0.523 | 0.011 |
| Restriction measures will eliminate the viral spread | 3.82 ± 1.59 | 0.709 | 0.000 | 0.017* | 0.011 | 0.002** | 0.028 |
| There should be no restrictions for religious operations | 1.62 ± 1.17 | 0.791 | 0.000 | 0.035* | 0.010 | 0.449 | 0.012 |
| Individual responsibility is more significant than state responsibility, toward coping with COVID-19 | 3.35 ± 1.71 | 0.495 | 0.000 | 0.162 | 0.007 | 0.783 | 0.009 |
| Herd immunity is an effective protection method | 2.08 ± 1.15 | 0.463 | 0.000 | 0.004** | 0.013 | 0.214 | 0.015 |
| I get regularly vaccinated | 2.72 ± 1.44 | 0.001* | 0.008 | <0.001*** | 0.023 | <0.001*** | 0.044 |
| I have adopted all the precaution measures against COVID-19 | 4.18 ± 0.85 | 0.002* | 0.007 | <0.001*** | 0.058 | 0.030* | 0.021 |

Agreement Likert scale: 1 (totally disagree) – 5 (totally agree); p: values from Kruskal Wallis test; , \*significant at p < 0.05, \*\*significant at p < 0.01, \*\*\*significant at p < 0.001; ε<sup>2</sup>: effect size; M: mean; SD: standard deviation

**Table 5.** Respondents confidence in institutions during the COVID-19 pandemic. Comparisons within study subgroups with the Kruskal Wallis nonparametric test (N=1396).

| Overall confidence levels and significance of sub-groups differences |  |  |  |  |  |  |  |
| --- | --- | --- | --- | --- | --- | --- | --- |
| Institutions | Confidence Level<br>(M ± SD) | Gender differences |  | Age differences |  | Educational level differences |  |
|  |  | p | ε<sup>2</sup> | p | ε<sup>2</sup> | p | ε<sup>2</sup> |
| World Health Organization | 3.85 ± 1.12 | 0.454 | 0.000 | <0.001*** | 0.032 | <0.001*** | 0.048 |
| Greek government | 3.14 ± 1.24 | 0.246 | 0.000 | 0.003* | 0.014 | 0.201 | 0.015 |
| Greek political parties | 1.90 ± 1.02 | 0.145 | 0.000 | 0.053 | 0.009 | 0.036* | 0.021 |
| Hellenic National Public Health Organization | 3.92 ± 1.07 | 0.842 | 0.000 | 0.605 | 0.003 | 0.007** | 0.025 |
| Panhellenic Medical Association | 3.66 ± 1.22 | 0.558 | 0.000 | 0.018* | 0.011 | 0.239 | 0.015 |
| Church | 1.52 ± 1.02 | 0.616 | 0.000 | 0.009** | 0.012 | 0.357 | 0.013 |
| Media | 2.09 ± 1.09 | 0.748 | 0.000 | <0.001*** | 0.020 | 0.337 | 0.013 |
| Social Networks | 1.89 ± 1.03 | 0.517 | 0.000 | 0.089 | 0.008 | 0.647 | 0.010 |
| Independent Scientists | 2.76 ± 1.22 | 0.791 | 0.000 | 0.195 | 0.006 | 0.006** | 0.025 |
| Internet sources | 2.32 ± 1.16 | 0.002* | 0.007 | 0.856 | 0.002 | 0.017* | 0.022 |

Confidence Likert scale: 1 (no/very low confidence) – 5 (very high confidence); p: values from Kruskal Wallis test; \*significant at $p < 0.05$ , \*\*significant at $p < 0.01$ , \*\*\*significant at $p < 0.001$ ; ε<sup>2</sup>: effect size; M: mean; SD: standard deviation

### 3.4 Principal Component Analysis

The KMO score was 0.77 and Bartlett’s test of sphericity was statistically significant (p < 0.001) supporting the application of factor analysis. The PCA results indicated five components with eigenvalues greater than 1.00, corresponding to a cumulative variance equal to 59.03% (Table 6).

**Table 6.** Eigenvalues and variance attributed to the principal components of the attitude and perception items identified by PCA.

| Component | Eigenvalue | % of Variance | Cumulative % |
| --- | --- | --- | --- |
| 1 | 3.862 | 23.40 | 23.40 |
| 2 | 2.290 | 15.47 | 38,87 |
| 3 | 1.835 | 8.74 | 47,61 |
| 4 | 1.242 | 5.91 | 53,52 |
| 5 | 1.158 | 5.51 | 59,03 |

The variables considered in the PCA and their factor loadings are shown in Table 7.

**Table 7.** Principal Component Analysis (PCA) of the attitude and perception survey items. Components loadings are the outcome of varimax rotation. Only loadings > 0.300 are shown.

|  | Components |  |  |  |  |
| --- | --- | --- | --- | --- | --- |
|  | 1 | 2 | 3 | 4 | 5 |
| Confidence in the Hellenic National Public Health Organization | 0.842 |  |  |  |  |
| Confidence in the WHO | 0.831 |  |  |  |  |
| Confidence in the Panhellenic Medical Association | 0.745 |  |  |  |  |
| Confidence in the Government | 0.610 |  |  |  | -0.346 |
| Violating the restriction measures is unethical |  | 0.709 |  |  |  |
| Restriction measures will eliminate the viral spread |  | 0.697 |  |  |  |
| Individual responsibility is more significant than state |  | 0.663 |  |  |  |
| Restriction measures had to be adopted regardless the economic impact |  | 0.632 |  |  |  |
| Confirmed COVID-19 cases that violate the restriction measures should pay at least partially for their treatment |  | 0.543 |  |  |  |
| The economic impacts of the lockdown will be tremendous |  | 0.416 |  |  | 0.378 |
| I have adopted all the precaution measures against COVID-19 |  |  |  |  |  |
| Confidence in social networks |  |  | 0.837 |  |  |
| Confidence in internet sources |  |  | 0.765 |  |  |
| Confidence in the media |  |  | 0.719 |  |  |
| Confidence in specialists |  |  | 0.592 |  |  |
| Confidence in Political Parties | 0.386 |  | 0.389 | 0.306 |  |
| Confidence in Church |  |  |  | 0.716 |  |
| There should be no restrictions for religious operations |  |  |  | 0.625 | 0.389 |
| The Public Healthcare System can cope with the COVID-19 outbreak |  |  |  | 0.486 | -0.343 |
| Restriction measures violate human rights |  |  |  |  | 0.706 |
| Herd immunity is an effective protection method |  |  |  |  | 0.388 |
| I get regularly vaccinated |  |  |  |  | -0.302 |

The five principal components corresponded to five attitude and perception patterns regarding COVID-19 identified in the study population. The first pattern was labeled “trust in institutions” and was correlated with high confidence levels in scientific and political institutions. The second pattern was labeled “trust in the restriction measures” and was characterized by the respondent’s perception of the necessity and the efficiency of the imposed measures, despite their impact on the economy. The third pattern was labeled “trust in media and the internet” and was correlated with high confidence levels in the respective factors. The fourth pattern was labeled “trust in traditional institutions” and was characterized by high confidence levels in the church and political parties. Finally, the fifth pattern was labeled “measures deniers” and was correlated with a high degree of agreement regarding the impact of the measures on the economy and on human rights and a negative perception of the vaccination, the ability of the Greek Public Healthcare System to cope with COVID-19 outbreak and the Government.

### 3.5 Association between attitude/perception patterns and knowledge scores

The measures of the linear model fit are shown in Table 8.

**Table 8.** Model fit measures for the association between the five principle components and the knowledge score.

| **Principal Component** | **R<sup>2</sup>** | **Adjusted R<sup>2</sup>** | **F** | **p** |
| --- | --- | --- | --- | --- |
| 1 | 0.0821 | 0.0689 | 6.19 | < .001 |
| 2 | 0.101 | 0.0816 | 5.16 | < .001 |
| 3 | 0.0413 | 0.0240 | 2.38 | < .001 |
| 4 | 0.0453 | 0.0315 | 3.28 | < .001 |
| 5 | 0.0972 | 0.0775 | 4.93 | < .001 |

R^2^ and adjusted R^2^ values indicated weak associations between the attitude and perception variables of each principal component and the knowledge score. Among them, the variables of principal component 2, named “trust in the restriction measures” fitted a slightly better with the linear model compared to the rest of the components (R^2^ = 0.101, adjusted R^2^ = 0.0816, p < 0.001). On the contrary, the variables loaded on principal component 3, “trust in media and the internet” had the lowest R^2^ and adjusted R^2^ values (0.0413 and 0.0240 respectively, p < 0.001).

## 4. DISCUSSION

This study assessed the level of knowledge of the Greek population about the COVID-19 etiology, prevention and treatment and their attitudes and perceptions regarding the restriction precaution measures and the trust they showed to scientific, political, and other institutions during the national lockdown period of the spring 2020 (March 23 - May 3). Assessing the acquisition of appropriate knowledge and the societal trust in institutions and authorities could provide an indication of the degree of preparedness of the community to effectively prevent and control the pandemic during these first months of the global health crisis.

The participants of the study had a moderate high knowledge about the disease etiology, diagnosis, transmission, symptoms, prevention, and treatment (overall median score: 74.8%). The age and educational level had stronger independent effects on the population’s knowledge. Higher educational level is a consistent indicator of higher knowledge scores regarding the pandemic in different studies. In contrast, the correlation between age and knowledge scores appears differentiated in different populations. For example, survey studies in Egypt [10], China [11], and Bangladesh [12] concluded that younger individuals tend to have higher knowledge scores. In these cases, the observed differences were attributed to an existing educational gap between people of different age, socioeconomic status, or place of residence (rural vs urban).

The accurate knowledge about the COVID-19 pandemic originates from the scientific authorities and media that reproduce the relevant information. On the other side, the lack of knowledge can be attributed to lack of interest, lack of access to information, or use of easily accessible but unreliable sources of information. An example of the latter are the social media which function as open access medical education providers, though in an uncontrolled online environment, thus raising concerns about the actual benefits and risks of people seeking information from these resources [13]. Misinformation has been characterized as the leading cause of confusion that hampers the appropriate attitudes toward the pandemic [14]. A potential underestimation of the severity of the disease and any misleading information about the significance of prevention can lead to individually and socially irresponsible and dangerous behaviors. Indeed, in a short time after the emergence of the disease in China, conspiracy theories and rumors regarding the origin of the virus and the efficiency of the prevention measures and treatments were globally spread through digital media accompanied by fearmongering and racist trends [13]. This is the reason why the WHO created the COVID-19 myth busters webpage focusing in the correction of circulating misinformation, thus fighting phenomena of mass panic [15].

Our results indicated that most of the participants adopted the imposed restriction and prevention measures, describing them as efficient, despite their significant impact on the economy. The participants demonstrated their confidence in scientific institutions and the Greek government. In contrast, the media, online social networks, and the Church did not attract the public’s trust. The level of education had the strongest effect on the attitudes and perceptions of the respondents, while gender was not a factor of significant differentiation. According to the results of pair-wise comparisons, more educated people showed a higher level of trust in the scientific organizations and a more positive attitude toward the efficiency of the restriction measures and the practice of vaccination. The latter seems to remain a controversial issue in Greece, as shown by both the overall score in the relevant item and the significant score differences between people of different gender, age and education.

An attitude against vaccination by a significant proportion of the Greek population had also been reported during the influenza A (H1N1) pandemic of 2009 [16]. Only 22.2% of the general population were then likely to accept vaccination, mainly because of uncertainty about the safety of the vaccine. The same barrier against vaccination has also been reported in other studies of the Greek [17] or other [18], [19] populations. The negative attitude toward vaccination against H1N1 in Greece was associated with female gender, ages 30-64 y.o. and perception of low risk of being infected or at risk because of infection. Compared to the above, in this study the anti-vaccination attitude observed during the first months of the pandemic was not correlated with gender, but with younger age and lower education.

The results regarding the trust of the respondents in institutions and authorities showed differences associated with the age and education level. More educated people expressed stronger trust in scientific institutions. Younger people trusted more the W.H.O. compared to older individuals. In contrast, older people showed more confidence in the media (television and radio) and the Church. The latter is reasonable since older people are expected to be closer to the religious culture of the country and familiar with the traditional means of communication, which provide them access to the information given by the authorities. Obviously, these findings reflect the public’s perceptions during a crisis period and they could be different if the pandemic was missing. However, there have been studies reporting almost unchanged public confidence in institutions before and during the COVID-19 crisis. The latter has been reported, for instance, for Sweden [20]. In order to have comparable data between countries, social and cultural parameters should be considered.

Historically, in periods of crisis, humans tend to shift toward religion. During the COVID-19 health crisis, Google searches for prayer increased by 50% according to data from 95 countries around the world. This increase was not related to socioeconomic factors, neither was a substitute for services in physical churches. Instead it was a means for coping with the current adversity [21]. For a significant proportion of the Greek population, the Church is not only a religious organization but a reflection of the Greek-Orthodox cultural and spiritual tradition. Thus, the values and fundamental principles of many Greeks were challenged because of the closure of churches and the abstention from rituals. All the above would look inconsistent to the low trust score for the Church recorded in the study, if the following clarification was not made: This score reflected the public confidence in the reliability of the Church authorities within the context of the health crisis and was not related to its significance as a component of the country’s culture. It seems that the respondents did not confuse their faith and tradition with the scientific knowledge required for coping with the disease.

Finally, in our study, Principal Component Analysis was implied to investigate the attitudinal barriers within the study population and potentially explain the trends revealed by the independent attitude and perception scores and the respective between-groups comparisons. In the same context, the five patterns identified were further assessed for their association with the knowledge scores. The two major patterns, “trust in scientific institutions” and “trust in the restriction measures” included variables correlated with a higher education level and exhibited a stronger association with higher knowledge scores. On the contrary, the “trust in traditional institutions” pattern showed an association with older ages and low knowledge scores, while the “measures deniers” was correlated with younger ages and lower education levels. These findings suggest that the continuum of attitudes and perceptions of the Greek population toward the COVID-19 pandemic is separated into areas, between which the barriers are set by the level of education, the age and the obtained knowledge about the disease. Attitudinal patterns indicating a higher level of individual and social responsibility and a positive attitude toward the scientific institutions and the implemented measures tended to be correlated with higher levels of education and knowledge score. On the contrary, negative attitudes were associated with lower education and younger ages.

In conclusion, the participants of this survey study showed a moderate high level of knowledge about COVID-19. However, knowledge was lower among males younger and less educated individuals. The participants had, in general, positive attitudes and perceptions toward the restriction measures and the scientific institutions. Age, education, and knowledge score were the factors defining the attitudinal patterns revealed. We conclude that education, as an overarching, long-term target, and public awareness of scientifically accurate information through multimedia reports, internet messages, campaigns etc. are the means for eliminating irresponsible and dangerous behaviors and for protecting individual and public health, social integrity, and prosperity in periods of health crises.

## Data Availability

The authors confirm that most of the data supporting the findings of this study are available within the article. The rest of them are available from the corresponding author, AA, upon reasonable request.
Due to the nature of this research, participants of this study did not agree for the raw data to be shared publicly.

## Limitations

Training material about COVID-19 pandemic was used for constructing the survey instrument, that was further validated through a pilot study. However, this was a cross-sectional study, conducted online once, at a particular time, during a global health crisis, and with the use of a convenience sample. Thus, generalizations were limited, and causality between variables could not be investigated.

## Authors’ Contribution

Andreas Anestis supervised the project, reviewed the literature, made the data analysis, and wrote the manuscript. Odysseas Lomvardeas conceived the presented idea, constructed, and administered the surveys and participated in data analysis and review of literature.

Nikolaos Papadakis gathered and analyzed the data and contributed to drafting the manuscript.

